# Estimating VO_2peak_ in 18-91 year-old adults: *Development and Validation of the FitMáx^©^ - Questionnaire*

**DOI:** 10.1101/2021.02.21.21249668

**Authors:** Renske Meijer, Martijn van Hooff, Nicole E. Papen-Botterhuis, Charlotte J.L. Molenaar, Marta Regis, Thomas Timmers, Lonneke V. van de Poll-Franse, Hans H.C.M. Savelberg, Goof Schep

**Affiliations:** Department of Sports and Exercise, Máxima Medical Centre, Veldhoven, the Netherlands; Department of Nutrition and Movement Sciences, NUTRIM School of Nutrition and Translational Research in Metabolism, Faculty of Health, Medicine and Life Sciences, Maastricht University, Maastricht, the Netherlands; Academy, Máxima Medical Centre, Veldhoven, the Netherlands; Department of Surgery, Máxima Medical Centre, Veldhoven, the Netherlands; Department of Mathematics and Computer Science, University Of Technology, Eindhoven, the Netherlands; Department of Research & Development, Rosmalen, the Netherlands; Department of Research and Development, Netherlands Comprehensive Cancer Organisation, Utrecht, Netherlands; Division of Psychosocial Research & Epidemiology, The Netherlands Cancer Institute, Amsterdam, Netherlands; Department of Medical and Clinical Psychology, Tilburg University, Tilburg, the Netherlands

## Abstract

**Objectives:** Cardiorespiratory fitness plays an essential role in health outcomes and quality of life. Objective assessment of cardiorespiratory fitness is costly, labour intensive and not widely available. Although patient-reported outcome measures estimate cardiorespiratory fitness more cost-efficiently, the current questionnaires lack accuracy. The aim of this study is to develop and validate the FitMáx©-questionnaire, a self-reported questionnaire to estimate cardiorespiratory fitness in healthcare.

**Methods:** We developed the FitMáx©-questionnaire, consisting of three questions assessing walking, stair climbing, and cycling capacity. A comparison on estimating VO_2peak_ was made with the Duke Activity Status Index (DASI), Veterans Specific Activity Questionnaire (VSAQ) and cardiopulmonary exercise testing as the gold standard. A total of 716 patients and athletes (520 men, 196 women) aged 18□91 performed a CPET in our hospital. We randomly selected 70% of the subjects to fit a linear regression model to estimate VO_2peak_ based on the FitMáx© scores. The remaining 30% of participants was used for validation of this model.

**Results:** The VO_2peak_ estimated by the FitMáx© strongly correlates with the VO_2peak_ measured objectively with CPET; r=0.95 (0.93□0.96) SEE=3.94 ml·kg^-1^·min^-1^. Bias between predicted and measured VO_2peak_ was 0.32 ml·kg^-1^·min^-1^ and the 95% limits of agreement were −8.11 □ 9.40 ml·kg^-1^·min^-1^. In our sample, the FitMáx scored superiorly on correlation and SEE compared with those from the DASI and VSAQ, r=0.80 (0.73□0.86) SEE=4.22 ml·kg^-1^·min^-1^ and r=0.88 (0.84□0.91) SEE=6.61 ml·kg^-1^·min^-1^, respectively.

**Conclusion:** FitMáx© is a valid and accessible questionnaire to estimate cardiorespiratory fitness expressed as VO_2peak_ and shows substantial improvement compared to currently used questionnaires.

**Key points:**

1. FitMáx© relies on three simple single-answer questions, which are recognizable for a large population, to accurately estimate cardiorespiratory fitness.
2. The FitMáx© is a self-reported instrument in which involvement of physicians, healthcare providers or other instrumentation is not necessary.
3. Cardiorespiratory fitness estimated by the FitMáx© may serve as an easily applicable measure in clinical and non-clinical settings.

## Introduction

Cardiorespiratory fitness (CRF), commonly defined as peak oxygen uptake (VO_2peak_), is considered a vital sign, and holds an essential role in health outcomes and quality of life.^1 2^ Low CRF is associated with all-cause mortality.^3 4^ Enhancement of CRF leads to improvements of quality of life, and diminishes disease-related symptoms, such as fatigue and depression. Increasing VO_2peak_ by only 3.5 ml·kg^-1^·min^-1^ has been associated with an 8□35% survival benefit in various study populations.^1 2 5^

Cardiorespiratory fitness can be objectively determined with cardiopulmonary exercise testing (CPET), which is increasingly used in clinical practice to diagnose heart and lung diseases, and determine causes of exercise limitation. One of the most important variables measured with CPET, is the maximum amount of energy obtained by aerobic metabolism, or VO_2peak_.^6-9^ The VO_2peak_, defined as the averaged peak oxygen uptake in the last 30 seconds of a CPET, is often used interchangeably with maximum oxygen uptake (VO_2max_). To determine VO_2max_, it is necessary to reach a plateau in VO_2_ uptake despite increasing work load. In clinical practice however, this plateau is often not reached, which makes VO_2peak_ the preferred measure to express CRF.^10^

Unfortunately, CPET is a costly, labour-intensive, not widely available test leading to limited applicability.^11 12^ An alternative way to assess CRF is use of self-reported questionnaires, such as the Duke Activity Status Index (DASI) and Veterans Specific Activity Questionnaire (VSAQ).^13 14^ The DASI, reached a correlation of r=0.81 with the VO_2peak_ measured by CPET when taken by a healthcare provider. In case of self-report however, DASI reached a correlation of r=0.58. Unfortunately, the SEE for the DASI questionnaire is not reported. The VSAQ estimates the metabolic equivalent of a task (MET) and reached a correlation of r=0.82 with maximal MET achieved on CPET and a standard error of the estimate (SEE) of 1.43 MET (5.0 ml·kg^-1^·min^-1^). A drawback of these questionnaires is that they use activities, such as basketball and skiing, which are not practiced globally.^15^

Therefore, the last author [GS], experienced in exercise testing, developed the FitMáx©-questionnaire, hereafter called FitMáx. This questionnaire consists of three single-answer multiple-choice questions regarding the maximum capacity for everyday activities: walking, stair climbing, and cycling. The aim of this study is to validate the FitMáx as a self-reported questionnaire to estimate CRF in combination with simple demographic information like age, sex, Body Mass Index (BMI).

## Methods

To evaluate the criterion validity of the FitMáx, cycle ergometer CPET was used as the gold standard measure for CRF. Additionally, FitMáx was compared with DASI and VSAQ in the same population to evaluate the construct validity. Data was collected prospectively from March 2018 until March 2020 in Máxima Medical Centre. This is a large Dutch non-academic teaching hospital with expertise in sports medicine and exercise physiology embedded in care for cardiac, pulmonary and oncologic patients.^16-18^ Approximately 20 CPETs are performed every week for diagnostic or scientific purposes, as well as part of (p)rehabilitation programs.

### Study population

Subjects aged ≥18 years, scheduled for CPET for medical reasons or as part of a health check, were sent the study information letter, informed consent form and the FitMáx, VSAQ and DASI questionnaires. Since CRF can change over time, we have chosen to include only subjects in the current study if documents were completed within 6 weeks (<42 days) prior to or after performing the CPET. To enable inclusion of patients with a cardiopulmonary exercise limitation or patients using beta-blockers who reached volitional maximal effort, we did not impose inclusion criteria for maximal exercise testing, such as respiratory exchange ratio (RER) >1.1 or 85% of age-predicted peak heart rate.^19^ A few cases (n=31) were excluded as the CPET was terminated submaximal by the physician due to e.g. uncontrolled arrhythmia or syncope. Moreover, if the FitMáx was incomplete, subjects were excluded. The authorized Medical Research Ethics Committee of Máxima Medical Centre issued a ‘non WMO acknowledgement’ for this study (reference number N18.051). The study was registered as NL8568 in the Netherlands Trial Register.

### Cardiopulmonary Exercise Testing

All CPETs were supervised by experienced sports physicians or a human movement scientist, and conducted according to international standards.^20^ Prior to CPET, maximum attainable workload (in Watts) was estimated based on patient characteristics and subjective physical exercise capacity. Based on this estimated maximum workload, a ramp protocol was used in which the subject was expected to reach the maximum load within approximately ten minutes. The physician who carried out the CPET was blinded for the results of the questionnaires which, if not blinded, could have biased the choice of exercise protocol. The tests were performed in a temperature-humidity controlled room. A 12-lead electrocardiogram was continuously recorded during rest, warm-up, exercise, and at least three minutes after maximal exercise (suction electrode KISS, GE Healthcare, Chicago, USA or Custo, CustoMed, GmbH, Ottobrunn, Germany). Gas exchange variables were measured breath-by-breath (Vyntus CPX, CareFusion, Hochberg, Germany or MetaLyzer 3B, Cortex, Leipzig, Germany).

### Questionnaires

The FitMáx consists of three single-answer questions about the maximum capacity of daily life activities that are frequently performed by the general Dutch population. Maximum walking capacity was chosen as a measure of CRF, since the distance walked during a six-minute walk test is strongly associated with VO_2peak_ in patients with severely reduced functional capacity.^21 22^ Maximum stair climbing capacity was chosen because previous studies indicate that the risk of perioperative pulmonary complications can be estimated with a stair climbing test.^23-25^ Lastly, maximum cycling capacity was used, since Dutch people often cycle in daily life and exercise testing is also performed on cycle ergometers to measure CRF.^26^ We tried to draft distinguishable answer options that are unequivocal, with steps as small as possible. The final FitMáx consists of a 0□13 scale for walking, 0-10 scale for stair climbing and 0-11 scale for cycling. The Dutch version of the FitMáx was translated into English, according to the translation procedure described in the guidelines for cross-cultural adaptation.^27^ The FitMáx is available in the supplementary file.

To assess the ability of subjects to estimate their maximum effort on the FitMáx, extra questions with a scale 1□10 were used for walking, stairclimbing and cycling capacity separately, in which 1 indicates “I cannot estimate properly” and 10 indicates “I can estimate properly”. These questions were added in a later phase of the study, and therefore results of n=167 participants are described. Beside the FitMáx, subjects were asked to complete the VSAQ from the beginning of the study.^14^ To expand the comparison with other physical fitness questionnaires, we also added the DASI to the validation study in April 2019.^13^ To enable direct comparison, results of all questionnaires were converted into VO_2peak_ in ml·kg^-1^·min^-1^, following the guidelines of these questionnaires.^13 14^

### Statistical analysis

Descriptive statistics for subjects’ characteristics are reported as mean and standard deviation (SD) in case of normal distribution, and median and interquartile range (IQR) otherwise. For continuous variables, unpaired Student’s t-tests were used to evaluate differences between groups. If the assumption of a normal distribution was not met, the Wilcoxon rank sum test was used instead. The Chi-squared test was used for categorical variables.

For the estimation of CRF via the FitMáx scores, linear regression was chosen after exploratory data analysis.

The steps of the FitMáx are ordered by definition. For the analysis then, each step of the FitMáx was replaced by the mean VO_2peak_ of all values from subjects’ self-reported scores. These values were then used as regression variable in the model, together with significantly associated dependent variables (age, sex and BMI).

To avoid overfitting, we created two subgroups: 70% of the subjects as training set and the remaining 30% as testing set. The random sample function in R was used for this.^28^

The training set was used to select the best-fitting linear regression model. We used stepwise regression to retain the variables that are most relevant for the prediction of CRF. We performed stepwise selection with 10-fold cross-validation with 100 repeats, retaining 20% of the data at each loop for validation. The residuals of the chosen model were examined on bias and heteroscedasticity (studentized Breusch-Pagan (Koenker-Bassett) test).^29 30^

The final model was validated on the testing set. The Pearson’s correlation coefficient was used to evaluate the linear relationship between the measured VO_2peak_ using CPET and the VO_2peak_ estimated by the FitMáx, VSAQ and DASI. Also, the coefficient of determination (R^2^) and SEE were calculated. Bland Altman plots were used to determine whether mean differences between estimated and measured VO_2peak_, with corresponding limits of agreement (LoA), are dependent on the size of the measured VO_2peak_ values.

The same methods were used to estimate the VO_2peak_ from the three FitMáx questions separately, and also from the FitMáx with walking and stairclimbing only. All computations were implemented in R (R-version 4.0).^28^ A p-value of <0.05 was considered statistically significant.

### Patient and public involvement

Before data collection started, the FitMáx was tested and improved in a pilot study with twenty patients via cognitive walkthrough. After this pilot study, minor adjustments were made and FitMáx was applied as a self-reported questionnaire in the current study.

## Results

### Patient characteristics

A total of 716 subjects (520 men and 196 women) who performed a CPET and completed the FitMáx were included for analysis (Figure 1). From the study population, 163 participants performed a CPET as part of a health check and 553 participants performed a CPET for medical reasons. Since the DASI was added in a later phase of the study, it was received by 524 subjects, and completed by 458 subjects. The VSAQ was not completed by 7 subjects. The training set consisted of 501 subjects and the testing set of 215 subjects.

**Figure 1.**
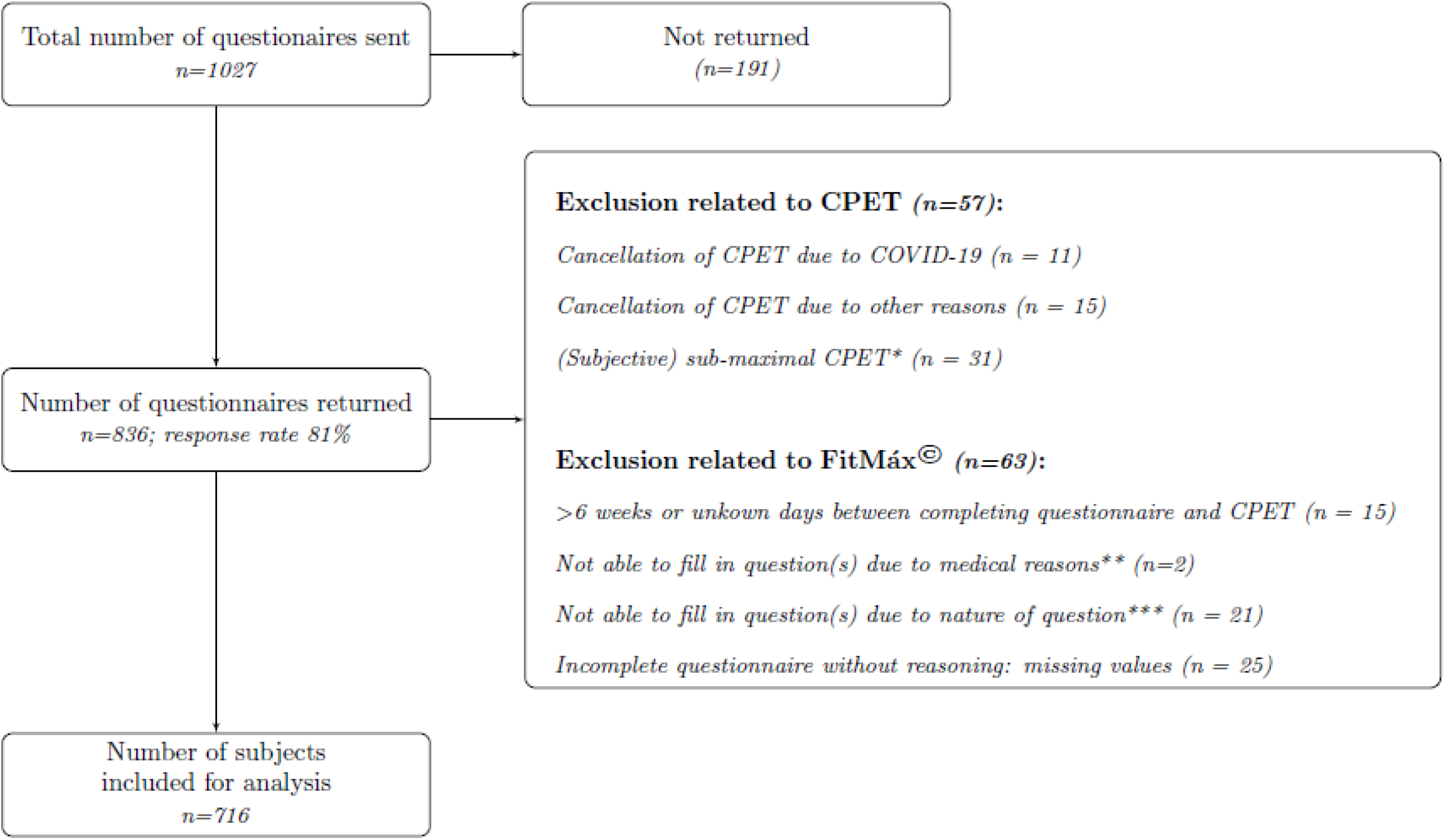
Flow diagram of participant selection Abbreviations: CPET, cardio-pulmonary exercise testing; COVID-19, coronavirus disease 2019; IC, informed consent. * Subjects with evidently submaximal performance on the CPET (i.e. not achieving volitional maximal effort), due to medical contraindications for maximal testing or measurement errors; ** Medical reasons were restrictions given by cardiologist & use of stair lift; *** e.g. never cycled before, use of electric bike

In the testing set, the subjects’ age ranged from 19-90 years with a VO_2peak_ from 9.6□71.4 ml·kg^-1^·min^-1^, whereas in the training group the subjects’ age ranged from 18-91 years with a VO_2peak_ from 7.5□67.2 ml·kg^-1^·min^-1^. Variables relevant for the interpretation of the CPET results include height, bodyweight, lung function, Global Initiative for Chronic Obstructive Lung Disease (GOLD) classification, use of beta-blockers and reason for CPET, and are presented in Table 1. No significant differences were found in the variables between testing and training set (Table 1). Median scores with inter quartile range of the patients’ ability to complete the FitMáx were 7 (8-9) for all three questions

**Table 1.** Participant Characteristics in the training and testing set, displayed separately

| Variable | Training Set (70%; n=501) |  | Testing Set (30%; n=215) |  |
| --- | --- | --- | --- | --- |
|  | Male | Female | Male | Female |
| n | 355 | 146 | 165 | 50 |
| *Anthropometrical data* |  |  |  |  |
| Age (yr) | 59.5 (49.0–67.6) | 60.6 (49.5–71.4) | 60.1 (51.2–68.8) | 57.3 (46.8–67.3) |
| Height (cm) | 180 (174–184) | 164 (160–170) | 181 (175–185) | 166 (161–172) |
| Weight (kg) | 82.4 (74.1–90.9) | 79.0 (61.5–81.6) | 82.5 (76.0–91.0) | 70.0 (60.5–79.0) |
| BMI (kg·m<sup>-2</sup>), | 25.4 (23.4–27.9) | 25.7 (22.6–30.0) | 25.7 (23.7–28.7) | 25.3 (21.5–27.7) |
| FEV<sub>1</sub> (L) | 3.6 (2.9–4.3) | 2.3 (1.8–3.0) | 3.5 (2.8–4.1) | 2.3 (1.8–3.0) |
| FVC (L) | 4.7 (3.8–5.5) | 3.0 (2.3–3.7) | 4.5 (3.8–5.2) | 3.0 (2.3–3.8) |
| COPD, GOLD classification |  |  |  |  |
| None | 324 (91.3%) | 118 (80.8%) | 149 (90.3%) | 44 (88.0%) |
| GOLD I | 10 (2.8%) | 2 (1.4%) | 1 (0.6%) | 2 (4.0%) |
| GOLD II | 15 (4.2%) | 18 (12.3%) | 8 (4.8%) | 4 (8.0%) |
| GOLD III | 5 (1.4%) | 6 (4.1%) | 5 (3.0%) | 0 (0%) |
| GOLD IV | 1 (0.3%) | 2 (1.4%) | 2 (1.2%) | 0 (0%) |
| Use of β-blocker |  |  |  |  |
| Yes, n (%) | 64 (18.0%) | 27 (18.5%) | 40 (24.2%) | 5 (10.0%) |
| No, n (%) | 291 (82.0%) | 119 (81.5%) | 125 (75.8%) | 45 (90.0%) |
| *CPET data* |  |  |  |  |
| Reason CPET/department |  |  |  |  |
| Health check, n (%) | 84 (23.7 %) | 20 (13.7 %) | 51 (30.9 %) | 8 (16.0 %) |
| Cardiac, n (%) | 176 (49.6 %) | 35 (24.0 %) | 72 (43.6 %) | 9 (18.0 %) |
| Pulmonary, n (%) | 66 (18.6 %) | 68 (46.6 %) | 28 (17.0 %) | 24 (48.0 %) |
| Oncologic, n (%) | 9 (2.5 %) | 15 (10.3 %) | 7 (4.2 %) | 4 (8.0 %) |
| Other reason, n (%) | 20 (5.6 %) | 8 (5.5 %) | 7 (4.2 %) | 5 (10.0 %) |
| Maximal workload (W) | 263 (145–347) | 114 (68–165) | 264 (133–350) | 106 (83–172) |
| Exercise time (min) | 9.5 (8.3–10.4) | 9.3 (7.8–10.6) | 9.3 (8.4–10.3) | 8.8 (7.4–10.1) |
| VO<sub>2peak</sub> (ml·kg<sup>-1</sup>·min<sup>-1</sup>) | 34.1 (21.0–43.9) | 20.6 (16.0–27.8) | 33.8 (19.2–43.2) | 19.9 (16.0–26.5) |
| VO<sub>2peak</sub> reference* (ml·kg<sup>-1</sup>·min<sup>-1</sup>) | 33.7 (29.6–37.6) | 22.2 (17.9–27.8) | 32.7 (28.6–36.9) | 23.5 (19.8–28.6) |
| HR<sub>peak</sub> (beat·min<sup>-1</sup>) | 157 (136–173) | 146 (123–171) | 160 (137–173) | 159 (128–173) |
| RER (VCO<sub>2</sub>/VO<sub>2</sub>) | 1.2 (1.1–1.2) | 1.1 (1.0–1.2) | 1.2 (1.1–1.2) | 1.1 (1.1–1.2) |
| *Questionnaire data* |  |  |  |  |
| VO<sub>2peak</sub> DASI (ml·kg<sup>-1</sup>·min<sup>-1</sup>) | 34.6 (25.0–34.6) | 24.5 (19.7–34.6) | 34.6 (22.1–34.6) | 28.0 (20.7–34.6) |
| VO<sub>2peak</sub> VSAQ (ml·kg<sup>-1</sup>·min<sup>-1</sup>) | 31.5 (17.0–40.1) | 16.6 (12.0–23.1) | 29.6 (15.6–38.7) | 18.6 (13.6–22.4) |
| VO<sub>2peak</sub> FitMáx (ml·kg<sup>-1</sup>·min<sup>-1</sup>) |  |  | 33.5 (19.5–41.4) | 21.9 (16.8–29.9) |
| Δ Time CPET and questionnaire (days) | 2 (0–8) | 0 (0–7) | 2 (0–8) | 1 (0–9) |

Results are displayed as n (%) or median (IQR).
Missing information, number of subjects: FEV<sub>1</sub>, 9; FVC, 9; Estimated VO<sub>2peak</sub> DASI, 258, VSAQ, 7.
Abbreviations: BMI, body mass index; cm, centimetres; COPD, chronic obstructive pulmonary disease; CPET, cardio-pulmonary exercise testing; DASI, duke activity status index; FEV<sub>1</sub>, forced expiratory volume in 1 second; FRIENDS, Fitness registry and importance of exercise national database\*; FVC, forced vital capacity; GOLD, Global initiative for chronic obstructive lung disease; HR, heart rate; kg, kilograms; kg·m<sup>-2</sup>, kilograms per square meter; L, litres; min, minutes; ml, millilitres; n, number of subjects; RER, respiratory exchange ratio; yr, years; VO<sub>2peak</sub>, peak oxygen uptake; VSAQ, veterans specific activity questionnaire; W, watts
\*The prediction model for VO<sub>2peak</sub> of the Fitness Registry and Importance of Exercise National Database (FRIENDS) is recommended by the American College of Sports Medicine as international reference value.<sup>7,38</sup>

### Development of the prediction model

Age, sex and BMI proved to be significantly associated with VO_2peak_ and were therefore included in the final model (p<0.05) (see Table 2). Homoscedasticity was not rejected by the studentized Breusch-Pagan (Koenker-Basset) test (p-value=0.45).

**Table 2.** Linear model FitMáx model fit. Standard error, t-values and p-values are reported for all variables included in the model.

| Term | Standard Error | Coefficient t-value | p-value |
| --- | --- | --- | --- |
| (Intercept) | 3.489 | 3.488 | <0.001 |
| Sex | 0.509 | -1.776 | <0.076 |
| Age | 0.046 | -0.069 | 0.945 |
| BMI | 0.051 | -7.356 | <0.001 |
| Walk | 0.093 | 5.383 | <0.001 |
| Stair climbing | 0.043 | 5.173 | <0.001 |
| Cycling | 0.040 | 12.829 | <0.001 |
| Walking*Age | 0.001 | -4.015 | <0.001 |

Abbreviations: BMI, Body Mass Index; Walk, maximum walking capacity score of the FitMáx; Stair climbing, maximum stair climbing capacity score of the FitMáx; Cycling, maximum cycling capacity score of the FitMáx
\* Sex is 0 for male and 1 for female, age in years with one decimal and BMI in kg·m<sup>-2</sup> with one decimal.

### Validation of the prediction model

Correlation of VO_2peak_ estimated by FitMáx with VO_2peak_ measured by CPET was higher r=0.95 (0.93□0.96) than the correlation of DASI r=0.80 (0.73□0.86) and VSAQ r=0.88 (0.84□0.91) (Figure 2a-c). Moreover, SEE and bias with LoA were smaller for the FitMáx and the coefficient of determination was higher compared to the same values for DASI and VSAQ corrected for the smaller complete subset of the DASI and VSAQ (Table 3). Bias of the FitMáx was 0.32 ml·kg^-1^·min^-1^, which is smaller than the same value for DASI (3.77 ml·kg^-1^·min^-1^) and VSAQ (3.70 ml·kg^-1^·min^-1^). Also, the results of predicting VO_2peak_ with the three FitMáx questions separately, and with the combination of the walking and stairclimbing capacity only are presented in Table 3. The estimated VO_2peak_ based on the walking and stairclimbing capacity reached a correlation of 0.92 (0.90□0.94) with VO_2peak_ measured by CPET. Although the values of correlation are comparable, SEE and LoA of the total FitMáx (including all three questions) are smaller than the combination of walking and stairclimbing only. Correlations of VO_2peak_ measured by CPET and the three FitMáx questions separately were r=0.89 (0.86□0.92) for walking, r=0.90 (0.87□0.92) for stairclimbing, and r=0.94 (0.92□0.95) for cycling.

**Table 3.**
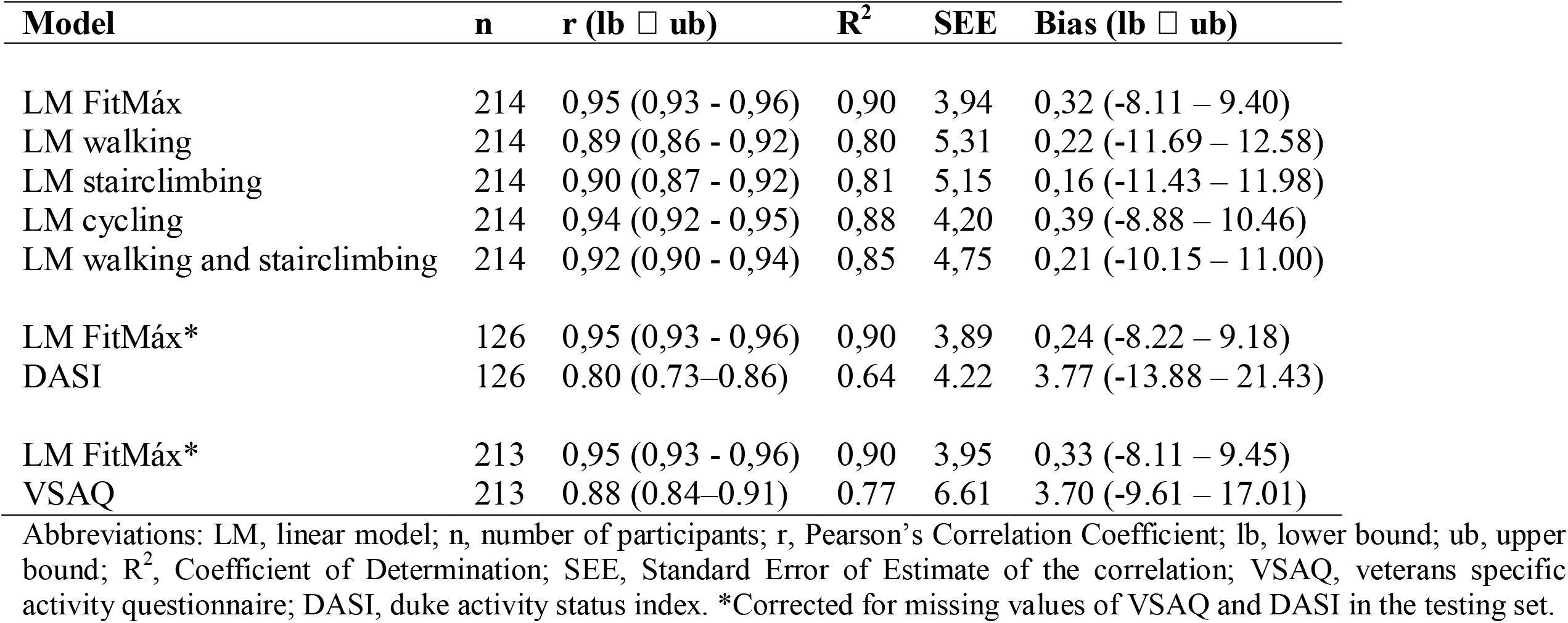
Statistics validation of the prediction model including walking, stair climbing and cycling capacity separately.

| Model | n | r (lb □ ub) | R<sup>2</sup> | SEE | Bias (lb □ ub) |
| --- | --- | --- | --- | --- | --- |
| LM FitMáx | 214 | 0,95 (0,93 - 0,96) | 0,90 | 3,94 | 0,32 (-8.11 – 9.40) |
| LM walking | 214 | 0,89 (0,86 - 0,92) | 0,80 | 5,31 | 0,22 (-11.69 – 12.58) |
| LM stairclimbing | 214 | 0,90 (0,87 - 0,92) | 0,81 | 5,15 | 0,16 (-11.43 – 11.98) |
| LM cycling | 214 | 0,94 (0,92 - 0,95) | 0,88 | 4,20 | 0,39 (-8.88 – 10.46) |
| LM walking and stairclimbing | 214 | 0,92 (0,90 - 0,94) | 0,85 | 4,75 | 0,21 (-10.15 – 11.00) |
| LM FitMáx* | 126 | 0,95 (0,93 - 0,96) | 0,90 | 3,89 | 0,24 (-8.22 – 9.18) |
| DASI | 126 | 0,80 (0,73–0,86) | 0,64 | 4.22 | 3.77 (-13.88 – 21.43) |
| LM FitMáx* | 213 | 0,95 (0,93 - 0,96) | 0,90 | 3,95 | 0,33 (-8.11 – 9.45) |
| VSAQ | 213 | 0,88 (0,84–0,91) | 0,77 | 6.61 | 3.70 (-9.61 – 17.01) |

Abbreviations: LM, linear model; n, number of participants; r, Pearson's Correlation Coefficient; lb, lower bound; ub, upper bound; R<sup>2</sup>, Coefficient of Determination; SEE, Standard Error of Estimate of the correlation; VSAQ, veterans specific activity questionnaire; DASI, duke activity status index. \*Corrected for missing values of VSAQ and DASI in the testing set.

**Figure 2.**
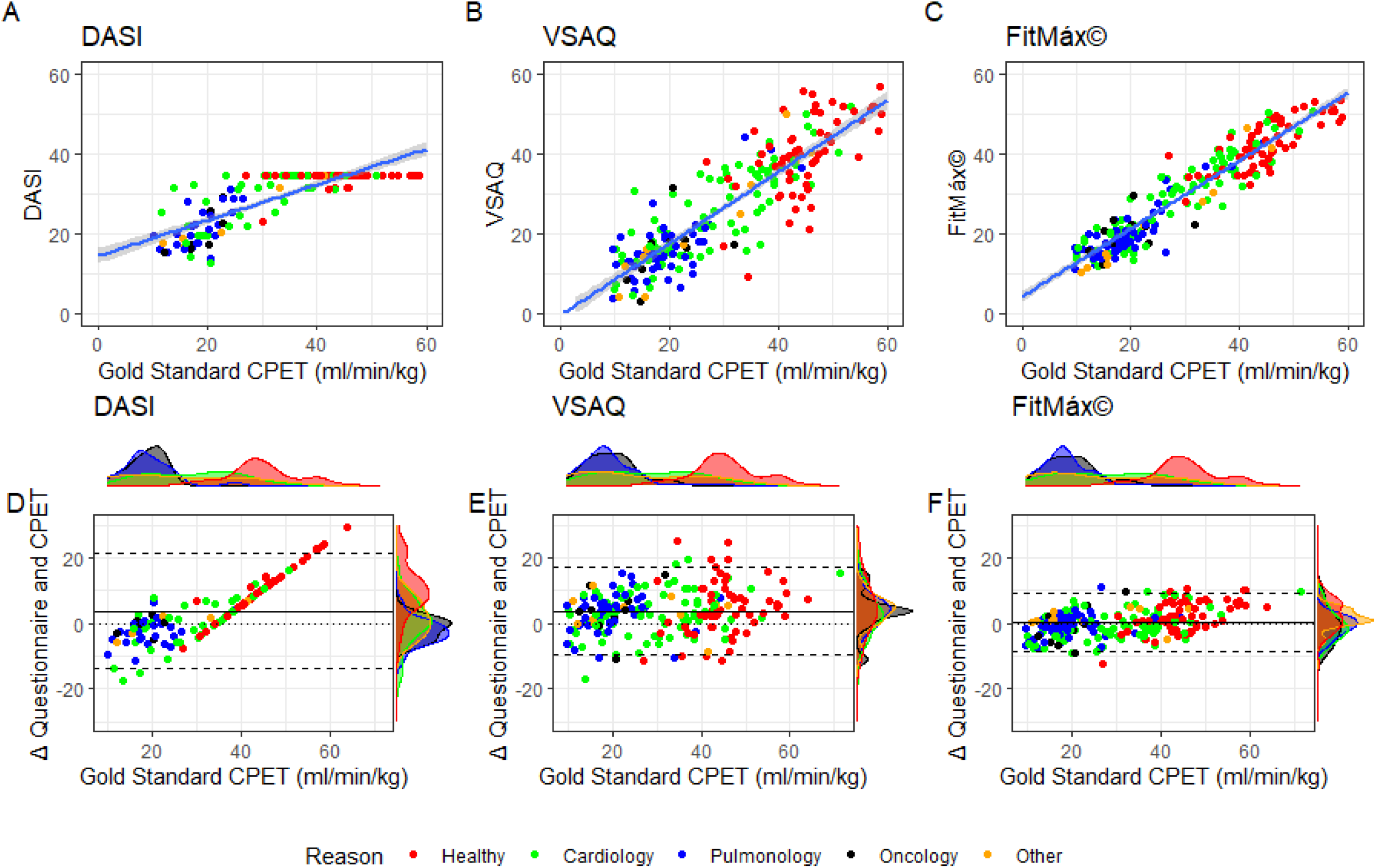
A-C) Scatterplots with identity line (i.e. perfect prediction) for FitMáx, DASI and VSAQ. The colours indicate the reason of the CPET visit; D-F)Bland Altman plots for DASI, VSAQ and FitMáx, above and on the right side of the axis histograms are plotted per reason of the CPET. The colours indicate the reason of the CPET visit. The dashed lines represent the limits of agreement, from −1.96 SD to +1.96 SD. The solid line represents bias and the dotted line represents the zero bias line. Abbreviations: CPET, cardio-pulmonary exercise testing; DASI, duke activity status index; min, minutes; ml, millilitres; kg, kilograms; r, Pearson’s Correlation Coefficient; VSAQ, veterans specific activity questionnaire.

Bland Altman analysis shows the agreement between measured and estimated VO_2peak_ by the FitMáx (−8.43□9.08 ml·kg^-1^·min^-1^) to be independent of the VO_2peak_ measured by CPET (Figure 2d-f). Density plots per indication of the CPET are displayed above and on the right side of the axis. The density plots on the y-axis of the FitMáx indicate that most of the results from the subjects are within the 95% LoA.

## Discussion

In the present study we developed a new questionnaire containing only three single-answer questions to estimate CRF. The VO_2peak_ estimated by the FitMáx showed a strong correlation (r=0.95) with the VO_2peak_ objectively measured by CPET. The validation was performed in a heterogeneous population of both healthy subjects and patients with variety in age and a wide range of CRF. Therefore, FitMáx proves to be applicable in young fit individuals, elderly and patients.

In our study population, the FitMax showed superior results compared with the DASI and VSAQ, which are questionnaires currently used in healthcare to estimate CRF. The FitMáx shows a good performance in a wide range of CRF and does not show floor or ceiling effects, in contrast to DASI.^31-33^ A possible explanation of these findings could be that the activities included in the VSAQ and DASI are more difficult to recognize for the Dutch population, compared to the activities included in the FitMáx.^15^

### Other instruments in literature

More recently, the CLINIMEX aerobic fitness questionnaire (C-AFQ) was developed, as a questionnaire-based prediction model for CRF.^34^ Despite a high correlation with CPET (r=0.91), the C-AFQ leaves considerable inaccuracy in the estimation of VO_2max_ (SEE= 5.39 ml·kg^-1^·min^-1^). Compared to the original C-AFQ study, correlation, SEE and bias with LoA are better for the FitMáx in the current study population.^34^ Moreover, an interview was used to complete the C-AFQ in the validation study, this could have led to a high correlation (r=0.91). This phenomenon is also seen in the validation of the DASI.^13^ However, to draw conclusions on the clinometric properties, the questionnaires should be compared within the same study population.

The prediction model of Bradshaw *et al*. is also questionnaire-based, and reached a correlation of r=0.91 with CPET and SEE=3.63 ml·kg^-1^·min^-1^ in the original research.^35^ The FitMáx showed a correlation of r=0.95 and a SEE=3.98 ml·kg^-1^·min^-1^ in our sample. The population in the research of Bradshaw *et al*. was small (100 participants) and had a relatively good CRF (mean VO_2max_ 39.96 ml·kg^-1^·min^-1^ ±9.54). It is unclear how the generalisability is affected by this specific study population.^35^

Finally, the HUNT study developed a prediction model to estimate VO_2peak_ that included age, physical activity, waist circumference, resting heart rate and peak heart rate.^36^ This approach is advised by the American Heart Association.^2^ However, applicability is limited to settings where such measurements are available. The correlation, SEE and bias reported in the HUNT study were worse than the ones found in the current FitMáx study, but results were obtained on the study-specific populations only.^36^ To draw conclusions, the two questionnaires should be compared on the same sample.

### Strengths of the current study

The strength of our study lies in the direct comparison of the performance of self-reported questionnaires to estimate VO_2peak_, with the same value measured by CPET as the gold standard, in a diverse population encompassing a wide range of age and CRF. Moreover, the physician involved in the CPET was blinded for the results of the questionnaires. If not blinded, this could have biased the choice of CPET protocol.

### Limitations of the current study

The study population consisted of mainly male subjects (70%). Analyses on sex showed that the FitMáx was able to estimate CRF accurately in both men and women. Nevertheless, to enhance the interpretability of the results for female subjects, more data should be collected. Moreover, no cross-cultural validations of VSAQ and DASI were available in Dutch, so the translation of the questionnaires was done by the researchers.

### Applicability of the FitMáx questionnaire

In the current study no physician or healthcare provider was involved in the completion of the questionnaire. FitMáx, DASI and VSAQ were evaluated as strictly self-reported measures for CRF. For the Dutch population, the three questions of the FitMáx are easy to relate to, and their sex, age, and BMI are usually known or easy to assess. For healthcare professionals, there is a well-known, single result of the questionnaire: VO_2peak_. For international use, the question about the maximum cycling capacity may be a limitation in the applicability of the FitMáx. However, estimated VO_2peak_ based on the FitMáx without the maximum cycling capacity still reached a correlation of r=0.92 (0.90□0.94) with measured VO_2peak_ by CPET. In the future we intend to add a question about daily activities to further improve the FitMáx. Despite the high correlation, the LoA are still relatively large indicating that discrepancies between patients’ self-reported and measured exercise capacity occur.^37^ Also the FitMáx estimates CRF, but does not diagnose the underlying limitation. FitMáx should not be considered as a full replacement for CPET, but rather a complementary tool to be used in settings where exercise testing is unavailable. Moreover, it may be used as a screening tool to detect patients with low CRF, who may benefit from an exercise intervention and/or more extensive diagnostic exercise testing.

### Implications for the future

To enhance the clinical applicability of the FitMáx, future studies will focus on its ability to monitor changes in CRF over time and on its comparison with other exercise tests, such as the steep ramp test and six-minute walk test.

To enable healthcare professionals and researchers in using the FitMáx questionnaire, we have developed an online platform (www.fitmaxquestionnaire.com) where we invite researchers to collaborate with us to further improve and validate the questionnaire in different settings. The online platform provides up-to-date information about the questionnaire and research projects. In addition to the online platform, a technical interface has been developed to implement the FitMáx questionnaire into third party applications. More information about our research group, hospital and FitMáx can be found on https://www.maximamc.com/fitmax.

## Conclusion

Cardiorespiratory fitness is of paramount importance in healthcare, given its substantial relation to both survival and quality of life. To tailor treatment and exercise interventions, it is important to measure cardiorespiratory fitness. The FitMáx consists of only three questions and is a simple tool to estimate cardiorespiratory fitness accurately.

## Supporting information

FitMax English Questionnaire

FitMax Dutch Questionnaire

## Data Availability

The datasets generated during and/or analyzed during the current study are available from the corresponding author on reasonable request.

## Acknowledgements

We would like to thank J. Dieleman, L. Bacas, A. den Bresser and J. Schellekens for their contribution to this research. Moreover, we would like to thanks J. van der Elsen,S. Bell, J. de Koning, T. Buscop and M. van Nieuwburg for helping with the forward backward translation method to provide the questionnaire in English.

