## Supplementary material for "Estimating VO_2peak_ in 18-91 year-old adults: *Development and Validation of the FitMáx^©^ - Questionnaire*": FitMax English Questionnaire

**List of questions regarding your fitness**

To determine how well you can walk/run, cycle and climb stairs, please answer the following questions by selecting **one** of the options below.

Choose the answer that best describes how well you can walk/run, cycle and climb stairs. This is what you can just maintain.

If you never walk/run, cycle or climb stairs, then try to imagine how it would be if you were to do this at this moment in time.

**1. Walking/running at this moment in time**

How well can you walk/run **at the moment**? Please select only **one** of the following options.

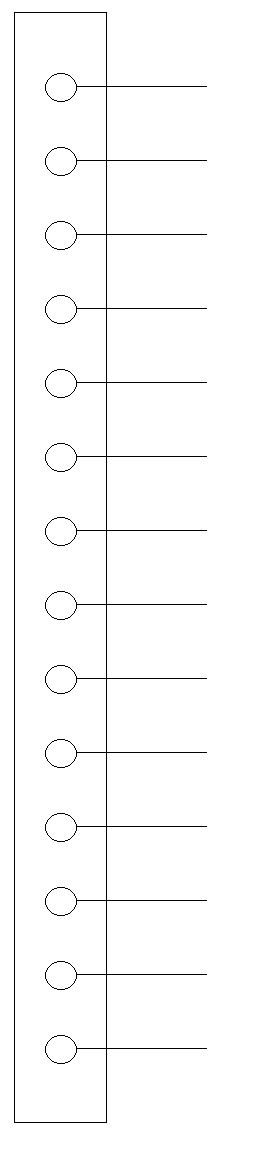

When I walk, I can’t carry on for long and have to **rest after 100** metres.

When I walk, I can’t carry on for long and have to **rest after 200** metres.

When I walk, I can’t carry on for long and have to **rest after 500** metres.

I can walk for **15 minutes** at a pace of **4 kilometres per hour (a gentle pace).**

I can walk for **30 minutes** at a pace of **4 kilometres per hour (a gentle pace).**

I can walk for **30 minutes** at a pace of **5 kilometres per hour (a normal pace).**

I can walk for **30 minutes** at a pace **of 6 kilometres per hour (brisk pace).**

I can **jog** for **30 minutes** at a pace of **7 kilometres per hour.**

I can **run** for **30 minutes** at a pace of **8-9 kilometres per hour.**

I can **run** for **30 minutes** at a pace of **10-11 kilometres per hour.**

I can **run** for **30 minutes** at a pace of **12-13 kilometres per hour.**

I can **run** for **30 minutes** at a pace of **14-15 kilometres per hour.**

I can **run** for **30 minutes** at a pace of **16-18 kilometres per hour.**

I can **run** for **30 minutes** at a pace **above 18 kilometres per hour.**

**2. Cycling at this moment in time**

How well can you cycle **at the moment**? Please select only **one** of the following options.

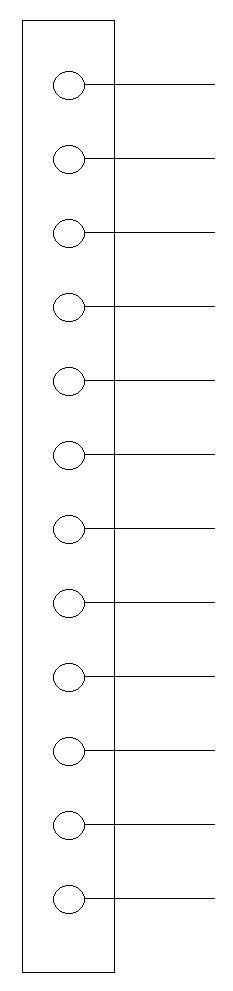

I am **not** **able** to cycle for **15** **minutes** on a flat road with no headwind on a normal bicycle.
te fietsen.

I can cycle for **15 minutes** on a flat road with no headwind on a normal bicycle, at a pace of **10-12 kilometres per hour (an idle pace).**

I can cycle for **15 minutes** on a flat road with no headwind on a normal bicycle at a pace of **13-15 kilometres per hour (a very slow pace).**

I can cycle for **30 minutes** on a flat road with no headwind on a normal bicycle, at a pace of **10-12 kilometres per hour (an idle pace).**

I can cycle for **30 minutes** on a flat road with no headwind on a normal bicycle, at a pace of **13-15 kilometres per hour (a very slow pace).**

I can cycle for **30 minutes** on a flat road with no headwind on a normal bicycle, at a pace of **16-18 kilometres per hour (a gentle pace).**

I can cycle for **30 minutes** on a flat road with no headwind on a normal bicycle, at a pace of **19-21 kilometres per hour (a normal pace).**

I can cycle for **30 minutes** on a flat road with no headwind on a normal bicycle, at a pace of **22-24 kilometres per hour (a brisk pace).**

I can cycle for **30 minutes** on a flat road with no headwind on a racing bike, at a pace of **25-30 kilometres per hour.**

I can cycle for **30 minutes** on a flat road with no headwind on a racing bike, at a pace of **30-35 kilometres per hour.**

I can cycle for **30 minutes** on a flat road with no headwind on a racing bike, at a pace of **35-40 kilometres per hour.**

I can cycle for **30 minutes** on a flat road with no headwind on a racing bike, at a pace **above 40 kilometres per hour.**

**3. Climbing stairs at this moment in time**

How well can you climb stairs **at the moment**? Please select only **one** of the following options.

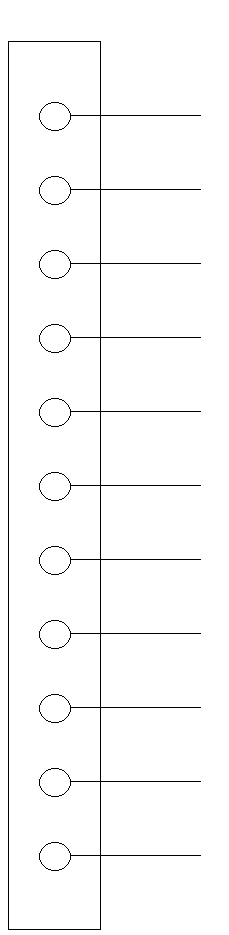

I am **not able** to walk up 1 flight of stairs without stopping.

I **am able** to climb **1 flight of stairs at a very slow pace** without stopping

I can walk up **2 flights of stairs at a very slow pace** without stopping.

I can run up **5 flights of stairs, taking two steps at a time,** without stopping.

I can **run** up **5 flights of stairs**, without stopping.

I can walk up **5 flights of stairs, taking two steps at a time**, without stopping.

I can walk up **5 flights of stairs at a normal pace** without stopping.

I can walk up **5 flights of stairs at a gentle pace** without stopping.

I can walk up **5 flights of stairs at a very slow pace** without stopping.

I can walk up **4 flights of stairs at a very slow pace** without stopping.

I can walk up **3 flights of stairs at a very slow pace** without stopping.

**4. Were you able to answer these questions easily?**

You have answered questions about how well you can walk/run, cycle or climb stairs **at the moment**. How well could you answer the questions? Circle the number that matches your experience with answering the questions.

| **1. Walk/run** | I could not estimate/answer correctly | 1 | 2 | 3 | 4 | 5 | 6 | 7 | 8 | 9 | 10 | I could estimate/answer correctly |
| --- | --- | --- | --- | --- | --- | --- | --- | --- | --- | --- | --- | --- |
| **2. Cycling** | I could not estimate/answer correctly | 1 | 2 | 3 | 4 | 5 | 6 | 7 | 8 | 9 | 10 | I could estimate/answer correctly |
| **3. Walking stairs** | I could not estimate/answer correctly | 1 | 2 | 3 | 4 | 5 | 6 | 7 | 8 | 9 | 10 | I could estimate/answer correctly |

**5. Circle one option which is most appropriate for you**

When, regardless of your fitness, you have stopped cycling, walking or climbing stairs because you have reached your limit:

- I am short of breath
- I feel pressure or pain on my chest
- My legs have given up
- I have problems with my joints
