## Supplementary material for "Estimating VO_2peak_ in 18-91 year-old adults: *Development and Validation of the FitMáx^©^ - Questionnaire*": FitMax Dutch Questionnaire

**Conditie vragenlijst**

Om te onderzoeken hoe goed u kan wandelen/lopen, fietsen en traplopen **op dit moment**, beantwoordt u de volgende vragen door per vraag **één** rondje aan te kruisen.

Kies hierbij het antwoord wat het beste omschrijft hoe goed u kan wandelen/lopen, fietsen en traplopen. Dit is wat u nog net kunt volhouden.

Als u nooit wandelt/loopt, fietst of trappen oploopt, probeer dan te bedenken hoe het zou zijn als u dat wel zou doen op dit moment.

**1. Wandelen/lopen op dit moment**

Hoe goed kan u wandelen/hardlopen **op dit moment?** Kruis **één** rondje aan.

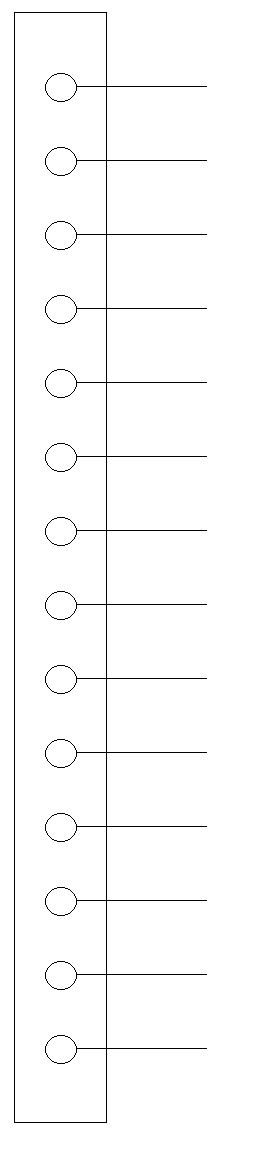

Als ik wandel houd ik dat niet lang vol en moet ik **even stoppen na 100** meter.

Als ik wandel houd ik dat niet lang vol en moet ik **even stoppen na 200** meter.

Als ik wandel houd ik dat niet lang vol en moet ik **even stoppen na 500** meter.

Ik kan **15 minuten** lang wandelen en volhouden met **4 kilometer per uur (rustig tempo).**

Ik kan **30 minuten** lang wandelen en volhouden op **4 kilometer per uur (rustig tempo).**

Ik kan **30 minuten** lang wandelen en volhouden op **5 kilometer per uur (normaal tempo).**

Ik kan **30 minuten** lang wandelen en volhouden met **6 kilometer per uur (stevig** **door wandelen).**

Ik kan **30 minuten** lang **joggen** en volhouden met **7 kilometer per uur.**

Ik kan **30 minuten** lang **hardlopen** en volhouden met **8-9 kilometer per uur.**

Ik kan **30 minuten** lang **hardlopen** en volhouden met **10-11 kilometer per uur.**

Ik kan **30 minuten** lang **hardlopen** en volhouden met **12-13 kilometer per uur.**

Ik kan **30 minuten** lang **hardlopen** en volhouden met **14-15 kilometer per uur.**

Ik kan **30 minuten** lang **hardlopen** en volhouden met **16-18 kilometer per uur.**

Ik kan **30 minuten** lang **hardlopen** en volhouden met **meer dan** **18 kilometer per uur.**

**2. Fietsen op dit moment**

Hoe goed kan u fietsen **op dit moment?** Kruis **één** rondje aan.

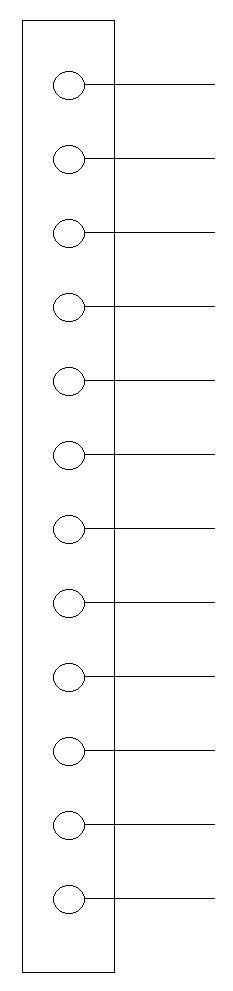

Ik kan **niet** **15 minuten** lang zonder tegenwind op een vlakke weg fietsen op een gewone fiets.
te fietsen.

Ik kan **15 minuten** lang zonder tegenwind op een vlakke weg fietsen op een gewone fiets en volhouden met **10-12 kilometer per uur (traag tempo).**

Ik kan **15 minuten** lang zonder tegenwind op een vlakke weg fietsen op een gewone fiets en volhouden met **13-15 kilometer per uur (heel rustig tempo).**

Ik kan **30 minuten** lang zonder tegenwind op een vlakke weg fietsen op een gewone fiets en volhouden met **10-12 kilometer per uur (traag tempo).**

Ik kan **30 minuten** lang zonder tegenwind op een vlakke weg fietsen op een gewone fiets en volhouden met **13-15 kilometer per uur (heel rustig tempo).**

Ik kan **30 minuten** lang zonder tegenwind op een vlakke weg fietsen op een gewone fiets en volhouden met **16-18 kilometer per uur (rustig tempo).**

Ik kan **30 minuten** lang zonder tegenwind op een vlakke weg fietsen op een gewone fiets en volhouden met **19-21 kilometer per uur (normaal tempo).**

Ik kan **30 minuten** lang zonder tegenwind op een vlakke weg fietsen op een gewone fiets en volhouden met **22-24 kilometer per uur (stevig doortrappen).**

Ik kan **30 minuten** lang zonder tegenwind op een vlakke weg fietsen op een racefiets en volhouden **met 25-30 kilometer per uur.**

Ik kan **30 minuten** lang zonder tegenwind op een vlakke weg fietsen op een racefiets en volhouden met **30-35 kilometer per uur.**

Ik kan **30 minuten** lang zonder tegenwind op een vlakke weg fietsen op een racefiets en volhouden met **35-40 kilometer per uur.**

Ik kan **30 minuten** lang zonder tegenwind op een vlakke weg fietsen op een racefiets en volhouden met **meer dan** **40 kilometer per uur.**

**3. Traplopen op dit moment**

Hoe goed kan u traplopen **op dit moment?** Kruis **één** rondje aan.

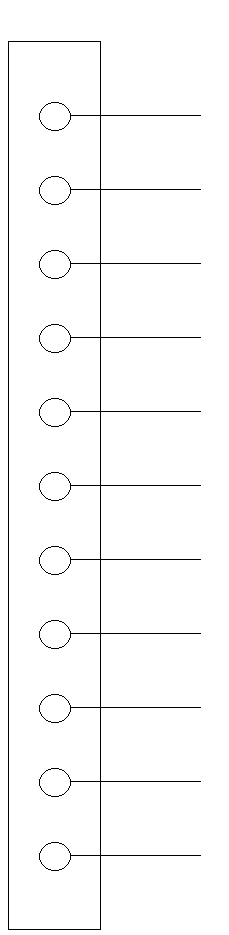

Het **lukt me niet** om **1 verdieping** trap te lopen zonder te stoppen.

Het lukt me **wel** om **1 verdieping heel rustig trap** te lopen zonder te stoppen

Het lukt me om **2 verdiepingen heel rustig** trap te lopen zonder te stoppen.

Het lukt me om **5 verdiepingen de trap op te rennen met twee treden tegelijk** zonder te stoppen.

Het lukt me om **5 verdiepingen de trap op te rennen** zonder te stoppen.

Het lukt me om **5 verdiepingen met twee treden tegelijk** trap te lopen zonder te stoppen.

Het lukt me om **5 verdiepingen met normaal tempo** trap te lopen zonder te stoppen.

Het lukt me om **5 verdiepingen rustig** trap te lopen zonder te stoppen.

Het lukt me om **5 verdiepingen heel rustig** trap te lopen zonder te stoppen.

Het lukt me om **4 verdiepingen heel rustig** trap te lopen zonder te stoppen.

Het lukt me om **3 verdiepingen heel rustig** trap te lopen zonder te stoppen.

**4. Kon u deze vragen makkelijk beantwoorden**

U heeft vragen beantwoord over hoe goed u kunt wandelen/lopen, fietsen en traplopen **op dit moment.** Hoe goed kon u de vragen beantwoorden? Zet een rondje om het getal wat het beste bij u past:

| **1. wandelen/ lopen** | Kan ik niet goed inschatten/ beantwoorden | 1 | 2 | 3 | 4 | 5 | 6 | 7 | 8 | 9 | 10 | Kan ik goed inschatten/ beantwoorden |
| --- | --- | --- | --- | --- | --- | --- | --- | --- | --- | --- | --- | --- |
| **2. fietsen** | Kan ik niet goed inschatten/ beantwoorden | 1 | 2 | 3 | 4 | 5 | 6 | 7 | 8 | 9 | 10 | Kan ik goed inschatten/ beantwoorden |
| **3. traplopen** | Kan ik niet goed inschatten/ beantwoorden | 1 | 2 | 3 | 4 | 5 | 6 | 7 | 8 | 9 | 10 | Kan ik goed inschatten/ beantwoorden |
